# Genetic discovery and risk prediction for type 1 diabetes in individuals without high-risk HLA-DR3/DR4 haplotypes

**DOI:** 10.1101/2023.11.11.23298405

**Authors:** Carolyn McGrail, Joshua Chiou, Ruth Elgamal, Amber M Luckett, Richard A Oram, Paola Benaglio, Kyle J Gaulton

**Affiliations:** Biomedical Sciences Graduate Program, UC San Diego, La Jolla, CA; University of Exeter College of Medicine and Health, Exeter, UK; Royal Devon University Healthcare NHS Foundation Trust, Exeter, UK; Department of Pediatrics, UC San Diego, La Jolla, CA

## Abstract

Over 10% of type 1 diabetes (T1D) cases do not have high-risk HLA-DR3 or DR4 haplotypes with distinct clinical features such as later onset and reduced insulin dependence. To identify genetic drivers of T1D in the absence of DR3/DR4, we performed association and fine-mapping analyses in 12,316 non-DR3/DR4 samples. Risk variants at the MHC and other loci genome-wide had heterogeneity in effects on T1D dependent on DR3/DR4, and non-DR3/DR4 T1D had evidence for a greater polygenic burden. T1D-assocated variants in non-DR3/DR4 were more enriched for loci, regulatory elements, and pathways for antigen presentation, innate immunity, and beta cells, and depleted in T cells, compared to DR3/DR4. Non-DR3/DR4 T1D cases were poorly classified based on an existing genetic risk score GRS2, and we created a new GRS which highly discriminated non-DR3/DR4 T1D from both non-diabetes and T2D. In total we identified heterogeneity in T1D genetic risk and disease mechanisms dependent on high-risk HLA haplotype and which enabled accurate classification of T1D across HLA background.

---

Type 1 Diabetes (T1D) is an autoimmune disease characterized by the destruction of insulin producing beta cells^1,2^ and which has complex etiology^2^. T1D is highly polygenic with over 90 known risk loci^3–8^12/2/23 9:34:00 PM and the largest genetic risk factors map to the Major Histocompatibility Complex (MHC) locus^7–9^ which encodes cell surface receptors that present antigenic peptides to CD4^+^ and CD8^+^ T cells^10^. Haplotypes of the class II HLA-DR and -DQ genes *DRB1*0301-DQA1*0501-DQB1*0201* (DR3) and *DRB1*04:01/02/04/05/08-DQA1*03:01-DQB1*03:02/04* (DR4)^7,11^ confer substantial risk of T1D^11^ and are detected in 90% of European T1D cases^12,13^. These high-risk DR3/DR4 haplotypes are thought to increase T1D risk by altering peptide binding to class II MHC and presentation of autoantigens to T cells^14^.

The remaining 10% of individuals of European ancestry that develop T1D without high-risk non-DR3/DR4 haplotypes have lower rates of T1D autoantibody positivity^15^ and later onset of disease with lower insulin dependence, which can be misdiagnosed as type 2 diabetes (T2D) or latent autoimmune diabetes in adults (LADA)^16–22^ leading to mismanagement and complications^16^. In addition, the preventative therapy Teplizumab is most effective in at-risk individuals who are DR4-positive and DR3-negative, and it is less established if it effectively delays onset in non-DR3/DR4 individuals^23^. Furthermore, existing genetic risk scores (GRS) for T1D heavily weigh DR3/DR4 haplotypes due to their large effect, and thus T1D in non-DR3/DR4 individuals may be poorly predicted by these scores. There is increasing evidence that adult onset T1D, which has a lower rate of DR3/DR4 haplotypes^24^, is more prevalent than previously estimated^25,26^, further underscoring the need to understand T1D in the absence of DR3 and DR4.

To address these gaps, we performed T1D association analyses in individuals without high-risk DR3/DR4 haplotypes (**Figure 1**). We obtained genotype data for 29,723 T1D and control individuals from five European ancestry cohorts (**Supplementary Table 1**), and imputed genotypes into 308M variants in TOPMed v2 and 56,310 variants in Michigan HLA reference panels^27,28^. We partitioned individuals by HLA-DR and -DQ haplotype resulting in 8,808 T1D and 8,199 controls with DR3/DR4 and 1,292 T1D and 11,424 controls without DR3/DR4 (**Supplementary Table 1**). We then tested for T1D association in the non-DR3/DR4 group, as well as the DR3/DR4 group for comparison. Six non-MHC loci *PTPN22, INS, IFIH1, PTPN2, IKZF4,* and *OSTN,* as well as the MHC locus, reached genome-wide significance (p<5×10^-8^) for T1D in non-DR3/DR4 (**Figure 1b, Supplementary Table 2**). Through conditional analysis (**see Methods**), we identified 12 additional signals at locus-wide significance (p<1×10^-5^), all of which were at the MHC locus (**Figure 1d, Supplementary Table 3**). For all signals we then generated 99% ‘credible sets’ of variants likely causal for T1D (**Figure 1c,e, Supplementary Table 4**).

**Fig. 1.**
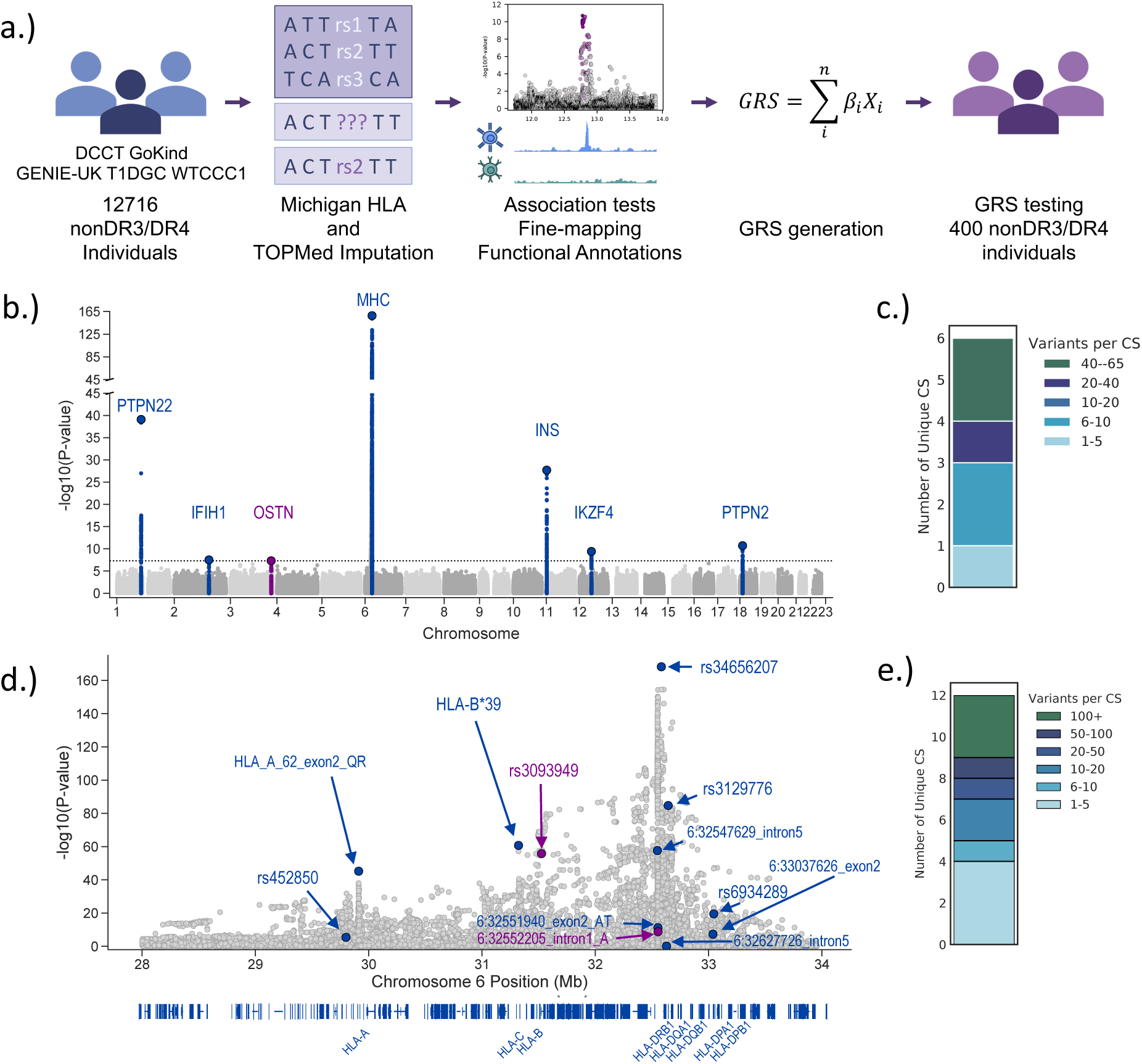
Genetic Discovery in Non-DR3/DR4 T1D. **a.** Overview of genetic discovery and risk prediction for T1D in non-DR3/DR4 individuals. **b.** Genome-wide T1D association (log_10_ *P* values from meta-analysis of *n*=12,316 samples) in individuals without DR3/DR4 haplotypes. Known T1D loci are colored blue and novel loci are colored purple. All loci are labeled with the nearest gene. **c.** Number of 99% credible set (CS) variants in fine-mapped non-HLA T1D risk signals. **d.** T1D association at the MHC locus (log_10_ *P* values from marginal association in meta-analysis of *n* = 12,316 samples). Known signals are colored blue and novel signals are colored purple. The location of class I and II HLA genes are shown on the bottom. **e.** Number of 99% credible set variants in fine-mapped MHC signals.

We compared the effects of risk variants at these loci on T1D between non-DR3/DR4 and DR3/DR4 using a Breslow-Day (BD) test (**see Methods**). Of the six significant non-HLA loci, five are known while one locus near *OSTN* is previously unreported **(Fig. 2a)**. At the *OSTN* locus, the lead variant had moderate effect in non-DR3/DR4 and limited effect in DR3/DR4 (b=0.4871, b= 0.0281, BD test p=1.29×10^-4^) **(Fig. 2b)**. We also observed a stronger effect in non-DR3/DR4 for lead variants at the *IFIHI* (b=-0.35, b=-0.091, p=3.88×10^-5^) and *PTPN2* (b=-0.37, b=-0.21, p=0.018) loci **(Fig. 2c),** both of which are implicated in beta cell function^29,30^, and this heterogeneity remained after conditioning on other known signals at these loci (**Supplementary Table 5**). We further examined lead variants of primary signals at 88 known T1D loci^3^, and observed heterogeneity (p<0.05) at 9 additional loci. At 5 loci there was evidence for larger effects in non-DR3/DR4 including *PRR15L*, *RAD51B, PRF1, PRKD2,* and 6q27, several of which have been implicated in both immune cells and beta cells. By comparison, 4 loci had evidence for smaller effect in non-DR3/DR4 14q32*, IL2RA*, *IL2*, and *CD69* **(Supplementary Fig. 1, Supplementary Table 6),** which are loci that largely affect T cell function.

**Fig. 2:**
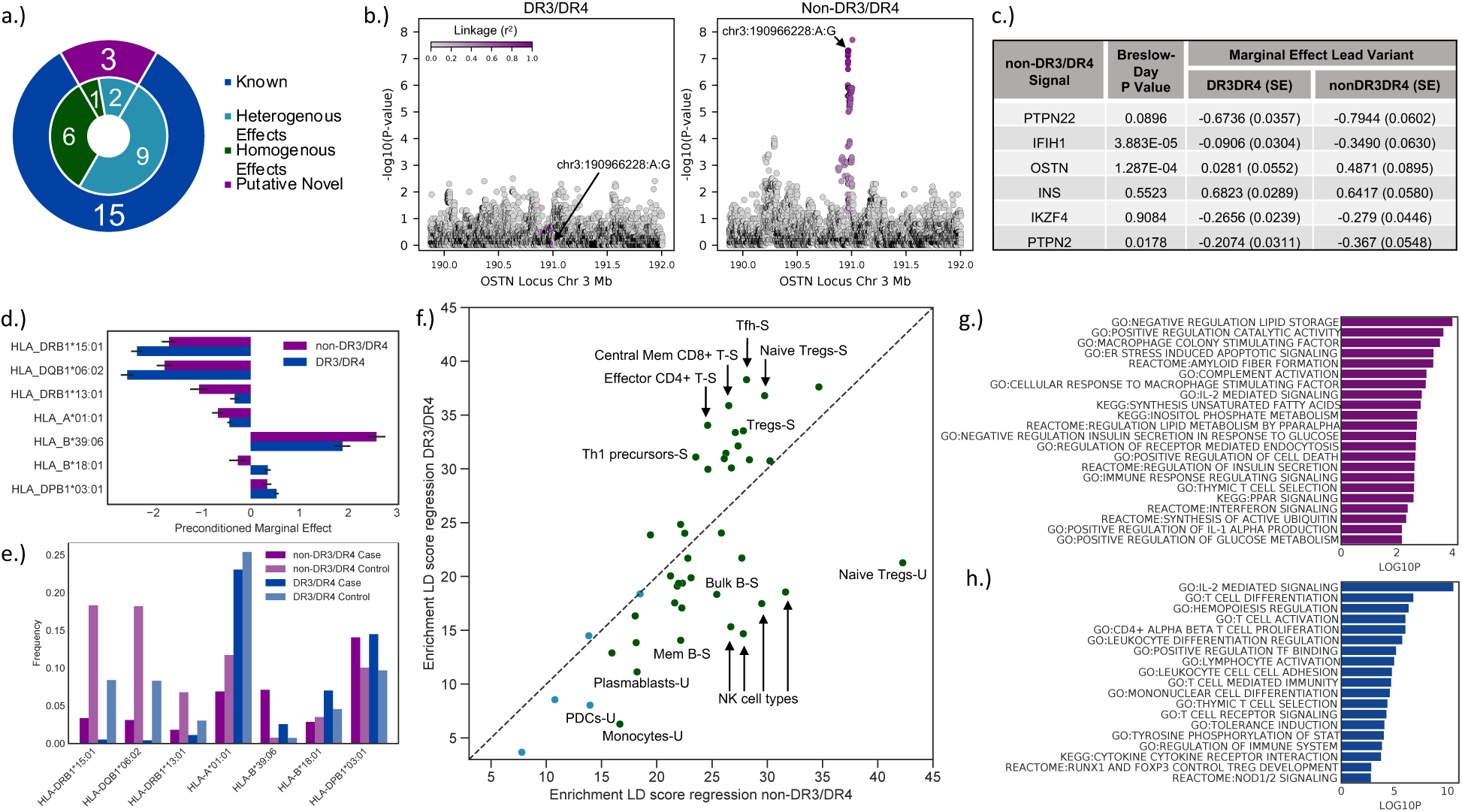
Heterogeneity of genetic and biological mechanisms in non-DR3/DR4 and DR3/DR4 T1D. **a**. Number of known and novel signals with heterogeneity in effect on T1D in non-DR3/DR4 and DR3/DR4 background. **b.** Locus plots of T1D association at the novel *OSTN* locus in DR3/DR4 (left) and non-DR3/DR4 (right). **c.** non-HLA loci with genome wide significant T1D association in non-DR3/DR4 analyses with effect sizes for lead variants in each group and difference in effect using interaction tests. **d.** T1D effect sizes of class I and II HLA alleles in each risk group after conditioning on 9 DR3 and DR4 risk alleles. **e.** T1D case and control frequencies of known HLA alleles in non-DR3/DR4 and DR3/DR4. **f.** Enrichment scores for T1D-associated variants in DR3/DR4 and non-DR3/DR4 in stimulated and unstimulated immune cell accessible chromatin using LD score regression. Dark green points are significant at FDR<0.10 in non-DR3/DR4 and DR3/DR4 while light blue points are FDR significant in only DR3/DR4. **g.** Gene pathway enrichment in non-DR3/DR4 T1D using MAGMA. **h.** Gene pathway enrichment in DR3/DR4 T1D using MAGMA.

We next determined whether individuals with T1D in the absence of HLA-DR3/DR4 may carry greater polygenic risk. We first determined whether known risk variants for T1D broadly had stronger effects in non-DR3/DR4 compared to DR3/DR4. An increased proportion (60.2%) of lead variants at 88 known T1D loci had stronger effect in non-DR3/DR4, although this was not significant (binomial p=0.069). We next determined whether non-HLA heritability differed between groups, and there was a small increase in estimated heritability for T1D in non-DR3/DR4 compared to DR3/DR4 (non-DR3/DR4 h^2^=0.2846, se=0.0795; DR3DR4 h^2^=0.2693, se=0.0428). Finally, there was a significant increase in the ability to distinguish T1D from controls in non-DR3/DR4 compared to DR3/DR4 (AUC=0.752, AUC=0.733, p=0.0238) using a risk score derived from 87 non-MHC lead variants in a recent T1D GWAS^3^. The ability to distinguish T1D from control in non-DR3/DR4 was further improved when using variant effects derived from the non-DR3/DR4 T1D GWAS (AUC=0.768, p=1.66×10^-5^). Together these results provide evidence for a larger polygenic contribution to T1D risk in the absence of high-risk DR3/DR4 haplotypes.

We next investigated whether variants within the MHC locus contribute to heterogeneity between non-DR3/DR4 and DR3/DR4 (**see Methods**). Of the 12 MHC signals identified in non-DR3/DR4 T1D, the lead variants of 8 signals exhibited heterogeneity between groups after conditioning on preceding signals from stepwise regression (**Fig. 2a, Supplementary Table 7**). Most signals were in linkage disequilibrium (LD) with known T1D-associated alleles, except for two apparent novel signals linked to HLA-DRB1*09:01-DQA1*03:01-DQB1*03:03 and B*44:02, respectively (**Fig. 2a, Supplementary Table 8**). Among a larger set of 40 known MHC T1D risk alleles, 22 two-digit class I and class II alleles had evidence for heterogeneity (p<0.05) (**see Methods, Fig. 2d,e, Supplementary Table 9**). In class I, HLA-B*39:06 had increased risk in non-DR3/DR4 (p=8.27×10^-3^), HLA-A*01:01 was more protective in non-DR3/DR4 (p=2.7×10^-4^), and HLA-B*18:01 had opposed effects in non-DR3/DR4 and DR3/DR4 (p=5.41×10^-6^) **(Fig. 2d,e)**. In class II, DRB1*15:01-DQB1*06:02 was more protective in DR3/DR4 (DRB1*15:01 p=8.187×10^-8^, DQB1*6:02 p=8.62×10^-7^) **(Fig. 2d,e)**, DRB1*13:01-DQB1*06:03 was more protective in non-DR3/DR4 (p=6.3×10^-3^) **(Fig. 2d,e)**, and DPB1*03:01 had increased risk in DR3/DR4 (p=0.035) **(Fig. 2d,e)**^31^. Together this reveals MHC alleles with heterogeneity in effect on T1D in a non-DR3/DR4 background.

As a higher proportion of individuals without DR3/DR4 haplotypes have later onset of disease, we next examined whether heterogeneity in T1D risk between non-DR3/DR4 and DR3/DR4 was consistent across age of onset. We compared the effects of non-DR3/DR4 signals in each individual cohort included in our meta-analysis, including the WTCCC and T1DGC cohorts where all cases had age of onset <17 (**Supplementary Table 1**). Among loci significant in non-DR3/DR4 that also exhibited heterogeneity with DR3/DR4, we observed no difference in T1D effects in WTCCC and T1DGC compared to other cohorts (Cochran Q p>0.05). (**Supplementary Table 2**, **Supplementary Fig. 2**). In addition, signals with heterogeneity between non-DR3/DR4 and DR3/DR4 were largely distinct from those with age-dependent association from a separate study^17^, and HLA-B*39:06 had larger effect in non-DR3/DR4 and was associated with younger age of onset (**Supplementary Fig. 1**). This supports that heterogeneity in non-DR3/DR4 appears distinct from T1D age of onset-dependent effects.

Given heterogeneity in effects at individual T1D loci in non-DR3/DR4 and DR3/DR4, we next determined if there were broader differences in T1D association between groups. We first tested T1D associated variants in each group for enrichment of candidate *cis*-regulatory elements (cCREs) in immune cell types using stratified LD score regression^32–34^. In the non-DR3/DR4 group, there was enrichment (FDR<0.10) of T1D associated variants in memory B-cells, bulk B cells, mature NK cells, and unstimulated T regulatory cells **(Fig. 2f, Supplementary Table 10)**. Dendritic cells and monocytes were also enriched in non-DR3/DR4 but did not pass multiple test correction. By comparison, the DR3/DR4 group was most enriched for cCREs in T cell populations including T follicular helper, naive T regulatory, central memory CD8^+^ T, and effector CD4^+^ T cells (**Supplementary Table 10)**. We next performed gene set enrichment in each group using MAGMA (**Supplementary Table 11)**. The strongest enrichments (p<0.05) in the non-DR3/DR4 included regulation of lipid storage, macrophage stimulating factor, stress-induced apoptotic signaling, and complement activation **(Fig. 2g)**. By comparison, DR3/DR4 was most enriched for T cell-related pathways including IL-2 signaling, and T cell differentiation, activation, and proliferation **(Fig. 2h)**. Collectively, this suggests a greater APC, innate immune and beta cell contribution, and lower T cell contribution, to T1D in non-DR3/DR4 compared to DR3/DR4.

Finally, we evaluated the ability of genetic risk scores (GRS) to predict T1D in non-DR3/DR4 individuals. Over 35% of non-DR3DR4 T1D cases were below the 5^th^ percentile for T1D in the published GRS2^35^, and only 7.5% were above the 50^th^ percentile used to classify T1D with high specificity. In line with this, GRS2 had less ability to distinguish non-DR3/DR4 T1D cases from all controls (AUC=0.764, **Fig. 3a,e**) as compared to all T1D cases (AUC=0.927)^35^. We thus generated a non-DR3/DR4-specific T1D GRS using the 18 risk signals identified in our non-DR3/DR4 GWAS (**see Methods, Supplementary Table 12**). This GRS improved discrimination of non-DR3/DR4 T1D from all controls compared to GRS2 (AUC=0.835; p=6.30×10^-15^) (**Fig. 3a,e,f**). We next generated a larger 100-variant non-DR3/DR4 T1D GRS by including all 82 non-MHC known T1D risk loci (**Methods, Supplementary Table 12**), which further discriminated T1D from all controls compared to GRS2 (AUC=0.867; p=7.48×10^-32^) (**Fig. 3b,e,g**). When both T1D cases and controls were subset to non-DR3/DR4 individuals, the 100-variant nonDR3/DR4 GRS again had improved discrimination of T1D over both the 18-variant GRS (AUC=0.894, AUC=0.871; p=5.70×10^-26^) (**Fig. 3c,f,g**) and GRS2 (AUC=0.878; p=0.0264) (**Fig. 3d,e,g**). We confirmed the ability of the 100-variant non-DR3/DR4 GRS to predict non-DR3/DR4 T1D using an independent test set of 100 T1D and 300 control non-DR3/DR4 samples excluded from the initial association analyses (AUC=0.887) (**Supplementary Fig. 3a,b**).

**Fig. 3.**
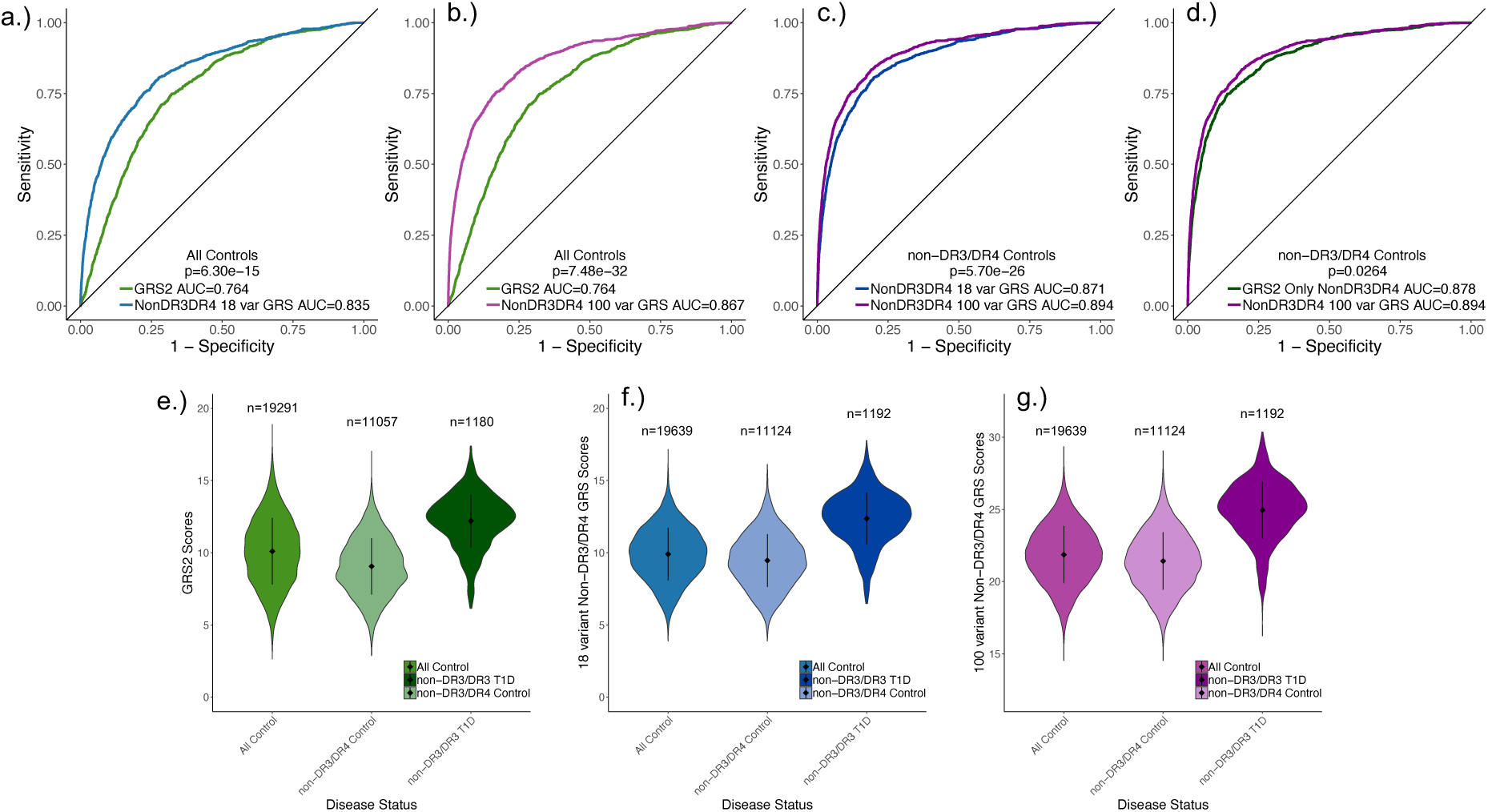
Genetic Risk Prediction in non-DR3/DR4 T1D. Receiver operating characteristic (ROC) curves showing the ability of genetic risk scores (GRS) to differentiate T1D from non-diabetes in the non-DR3/DR4 population and corresponding violin plots. The AUCs are shown for each GRS and the p-values comparing predictive ability of GRS are calculated using the deLong test. **a.** GRS2 compared to the 18 variant non-DR3/DR4 GRS in non-DR3/DR4 T1D vs. all controls. **b.** GRS2 compared to the 100-variant non-DR3/DR4 T1D GRS in non-DR3/DR4 T1D vs. all controls. **c.** The 18-variant non-DR3/DR4 T1D GRS compared to the 100-variant non-DR3/DR4 T1D GRS in non-DR3/DR4 T1D and controls. **d.** GRS2 subset to non-DR3/DR4 T1D and controls compared to the 100-variant non-DR3/DR4 T1D GRS in non-DR3/DR4 T1D and controls. Violin plots for risk scores in all controls, non-DR3/DR4 controls, and T1D cases depicted for **e.** GRS2 **f.** 18-variant non-DR3/DR4 GRS and **g.** 100-variant non-DR3/DR4 GRS.

Since non-DR3/DR4 T1D individuals tend to have later onset and a lower dependence on insulin therapy leading to potential misdiagnosis as T2D^16,17,24,36^, we next evaluated the ability of a GRS to differentiate T1D from T2D using the WTCCC1 cohort (**Fig. 4**). Using GRS2 there was limited ability to distinguish non-DR3/DR4 T1D from all T2D (AUC=0.759) (**Fig. 4a,e**). In contrast, our non-DR3/DR4 GRS strongly discriminated non-DR3/DR4 T1D from T2D and was a significant improvement over GRS2 (18-variant AUC=0.890, p=1.51×10^-32^; 100-variant AUC=0.907, p=4.94×10^-44^) **(Fig. 4a,b,e,f,g)**.

**Fig. 4.**
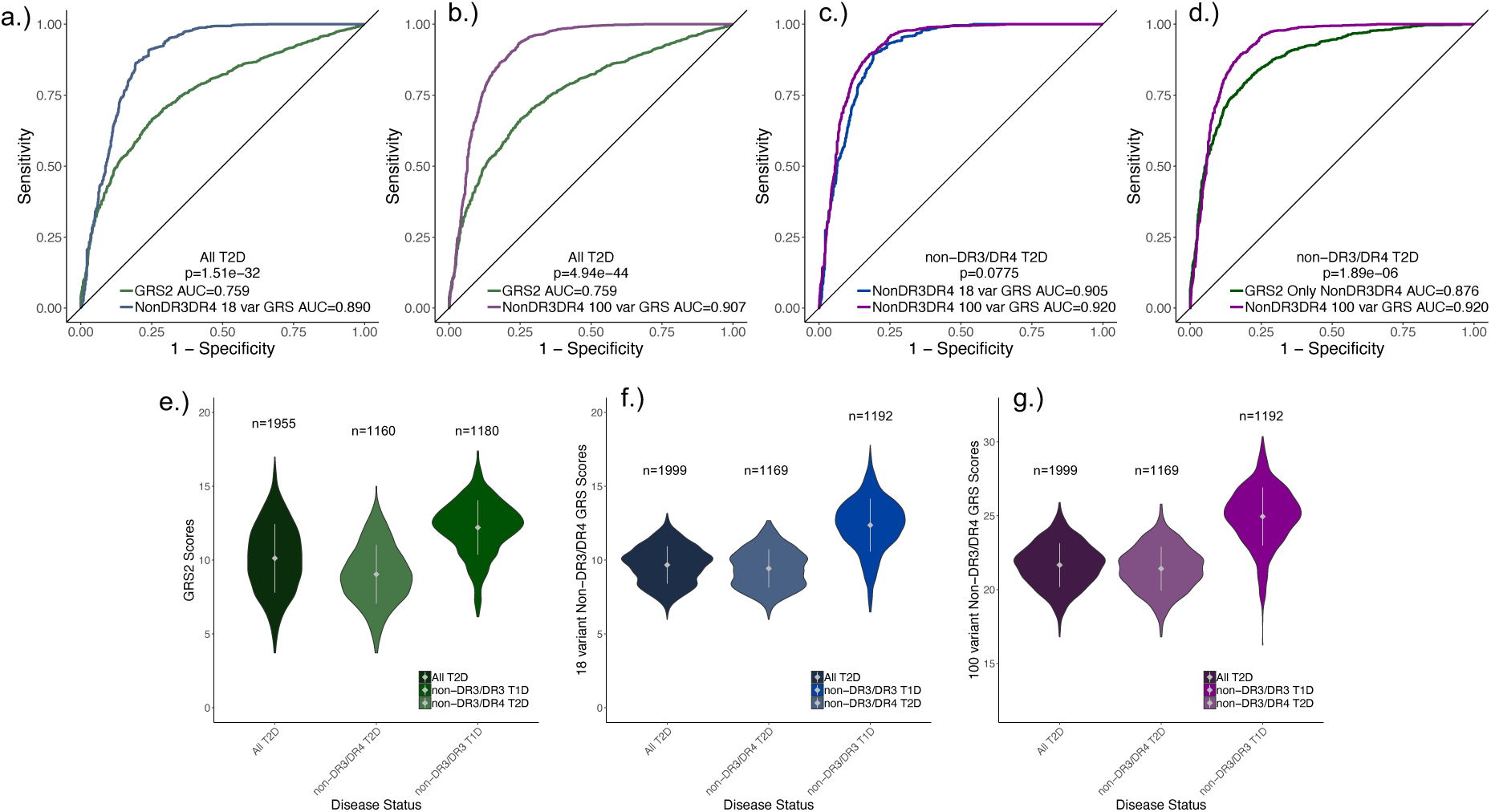
Genetic Risk Prediction of non-DR3/DR4 T1D and T2D. ROC curves showing the ability of GRS to differentiate T1D from T2D in the non-DR3/DR4 population and corresponding violin plots. The AUCs are shown for each GRS and p-value for difference in prediction of T1D from T2D is calculated using the deLong test. **a.** GRS2 compared to the 18 variant non-DR3/DR4 GRS in non-DR3/DR4 T1D vs. all T2D. **b.** GRS2 compared to the 100-variant non-DR3/DR4 T1D GRS in non-DR3/DR4 T1D vs. all T2D. **c.** The 18-variant non-DR3/DR4 T1D GRS compared to the 100-variant non-DR3/DR4 T1D GRS in non-DR3/DR4 T1D and T2D. **d.** GRS2 subset to non-DR3/DR4 T1D and T2D compared to the 100-variant non-DR3/DR4 T1D GRS in non-DR3/DR4 T1D and T2D. Violin plots for risk scores in all T2D, non-DR3/DR4 T2D, and T1D cases depicted for **e.** GRS2 **f.** 18-variant non-DR3/DR4 GRS and **g.** 100-variant non-DR3/DR4 GRS.

When both T1D and T2D cases were subset to non-DR3/DR4 individuals, the 100-variant non-DR3/DR4 GRS improved on both the 18-variant GRS (AUC=0.920, AUC=0.905, p=0.0775) (**Fig. 4c,f,g**) and GRS2 (AUC=0.876, p=1.89×10^-6^) (**Fig. 4d,e,g**). The predictions were consistent when utilizing an independent set of 100 T1D samples (AUC=0.906) (**Supplementary Fig. 3c,d**). The ability to distinguish non-DR3/DR4 T1D from T2D appears largely driven by the improved estimation of effects at the MHC locus (T1D vs T2D HLA AUC=0.885), as the extensive HLA combinations in GRS2 still do not enable differentiating non-DR3DR4 T1D from T2D (T1D vs T2D GRS2 HLA AUC=0.698).

We finally determined the diagnostic value of a GRS for T1D in a non-DR3/DR4 background. Non-DR3/DR4 individuals with T1D on the published GRS2 scale do not meet the minimum requirement for diagnostic feasibility at a Youden index of 0.50 (max Youden index=0.427). When T1D GRS2 is subset to only non-DR3/DR4 and re-scaled (**Table 1**), a score of 12.40 at the 50th percentile of T1D has a sensitivity of 50% and specificity of 95.4% and meets diagnostic criteria (max Youden index=0.609) at the 25^th^ T1D percentile. When comparing T1D to T2D using GRS2, there was limited ability to distinguish T1D from T2D (max Youden index=0.414). By comparison, the 18 variant non-DR3/DR4 GRS has a score of 12.50 at the 50th percentile for T1D with a sensitivity of 50.0% and specificity of 95.2% (**Table 1**) and a peak Youden index of 0.607 at a score threshold of 11.05. Similarly, in the 100-variant non-DR3/DR4 GRS, the 50^th^ percentile of T1D has a sensitivity of 50.0% and specificity of 96.9% and a peak Youden index of 0.644 at a score threshold of 23.21 (**Table 1**). There was also diagnostic value in the 100-variant non-DR3/DR4 GRS in differentiating non-DR3/DR4 T1D from T2D (max Youden index=0.687) and all controls (max Youden index=0.585). Overall, our results provide a GRS with clinical value in distinguishing non-DR3/DR4 T1D from non-diabetes and T2D.

**Table 1:** Sensitivity, Specificity, Youden Index, and scales for GRS cutoff values in non-DR3/DR4 only GRS2, non-DR3/DR4 18 variant GRS and non-DR3/DR4 100 variant GRS.

| **non-DR3/DR4 only GRS2** | **T1D Centile** | **Non-Disease Centile** | **Sensitivity (%)** | **Specificity (%)** | **Youden Index** |
| --- | --- | --- | --- | --- | --- |
| 8.86 | 5 | 47.6 | 95.00 | 47.63 | 0.4263 |
| 10.33 | 15 | 73.7 | 85.00 | 73.72 | 0.5872 |
| 11.20 | 25 | 85.3 | 75.00 | 85.35 | 0.6035 |
| 11.77 | 35 | 91.3 | 65.00 | 91.27 | 0.5627 |
| 12.40 | 50 | 95.4 | 50.00 | 95.41 | 0.4541 |
| 13.39 | 75 | 98.9 | 25.00 | 98.85 | 0.2385 |
| 14.85 | 95 | 99.9 | 5.000 | 99.95 | 0.0978 |
| **18-variant non-DR3/DR4 GRS** | **T1D Centile** | **Non-Disease Centile** | **Sensitivity (%)** | **Specificity (%)** | **Youden Index** |
| 9.066980 | 5 | 42.0 | 94.97 | 41.95 | 0.3691 |
| 10.585440 | 15 | 72.5 | 84.98 | 72.60 | 0.5758 |
| 11.358825 | 25 | 84.5 | 75.00 | 84.53 | 0.5953 |
| 11.850500 | 35 | 90.3 | 65.02 | 90.26 | 0.5527 |
| 12.497350 | 50 | 95.2 | 50.00 | 95.20 | 0.4519 |
| 13.553850 | 75 | 98.8 | 25.00 | 98.78 | 0.2377 |
| 15.080665 | 95 | 99.9 | 5.034 | 99.87 | 0.04908 |
| **100-variant non-DR3/DR4 GRS** | **T1D Centile** | **Non-Disease Centile** | **Sensitivity (%)** | **Specificity (%)** | **Youden Index** |
| 21.254775 | 5 | 46.9 | 94.97 | 46.90 | 0.4187 |
| 23.057930 | 15 | 78.7 | 84.98 | 78.68 | 0.6366 |
| 23.843050 | 25 | 88.4 | 75.00 | 88.41 | 0.6341 |
| 24.434680 | 35 | 93.6 | 65.02 | 93.56 | 0.5858 |
| 25.108450 | 50 | 96.9 | 50.00 | 96.90 | 0.4689 |
| 26.283800 | 75 | 99.3 | 25.00 | 99.33 | 0.2433 |
| 27.820655 | 95 | 99.9 | 5.034 | 99.96 | 0.0450 |

## Discussion

We observed heterogeneity in T1D genetic risk dependent on high-risk DR3/DR4 haplotypes at class I and class II MHC alleles as well as other loci genome wide. Multiple genes at loci with larger effects on T1D in non-DR3/DR4 such as *PTPN2, IFIH1, PRR15L,* and *RAD51B* are implicated in inflammatory signaling^37,38^ and survival^29,39,40^ in immune cells and beta cells. We also identified a novel T1D locus in non-DR3/DR4 near *OSTN,* which has been linked to diabetic cardiomyopathy in mouse models^41^. Conversely, many loci with reduced effect in non-DR3/DR4 such as *IL2RA*, *IL2* and *CD69* impact T cell function and activation^42,43^. More broadly, there were distinct patterns of enrichment in non-DR3/DR4 T1D where genomic annotations and molecular pathways related to B cells, NK cells, and beta cells were more enriched, and T cells less enriched, compared to DR3/DR4. These results collectively suggest that mechanisms of T1D in a non-DR3/DR4 background may depend more on inflammation and beta cell dysfunction than T cell activation^16–20^. Given modest evidence for a higher polygenic burden in T1D in a non-DR3/DR4 background, individuals without this high-risk factor may require additional polygenic risk to lead them to develop T1D.

Based on previous GRS percentile scales, many non-DR3/DR4 individuals with T1D would not have been predicted to develop T1D. The GRS reported here enables accurate discrimination of T1D from non-diabetes and T2D in non-DR3/DR4 individuals. To improve accessibility, the variants in the non-DR3/DR4 GRS are derived from publicly available imputation panels enabling the GRS to be more easily calculated from genotyping array data. While the subset of individuals that develop T1D without DR3/DR4 exhibit later onset and lower insulin dependence, they have similar risk profiles to the majority of T1D cases and not to T2D. The accurate prediction of T1D in the non-DR3/DR4 population in this study will therefore help avoid misdiagnosis of T2D and prevention of ketoacidosis and future complications. While non-DR3/DR4 T1D background has generally later onset, it can occur at any age and our results suggest that the effects of variants on T1D risk in a non-DR3/DR4 background are generally consistent across age of onset. As this study was conducted in individuals of European ancestry, similar studies of T1D in other ancestries may help improve risk prediction of T1D in these populations. Furthermore, the GRS reported here may have value in discriminating T1D in populations where high-risk haplotypes such as DR3/DR4 are uncommon.

The ability to distinguish at-risk individuals is also critical for determining eligibility for clinical trials and therapies. The preventative therapy Teplizumab modulates T cell activity and is most efficacious in individuals with HLA-DR4^23^, and several clinical trials aiming to preserve beta cell function at onset preferentially recruit DR3/DR4 individuals^44,45^. Given evidence for differences in disease mechanisms, in particular a less prominent contribution from T cells, alternate therapies may be needed to prevent T1D in non-DR3/DR4 individuals. In addition, as autoantibodies are seen at lower rates in non-DR3/DR4 T1D, additional biomarkers are needed for this group^15^.

Overall our findings are in line with a growing body of literature ^46–49^ which support that T1D is a heterogeneous disease consisting of sub-types with distinct pathophysiological features. More broadly, stratifying by high-risk genetic background may be an effective strategy to discover genetic and mechanistic heterogeneity in other complex diseases.

## ONLINE METHODS

### Research subjects and genotype imputation

We compiled genotype data from 10,100 T1D and 19,623 control individuals of European ancestry from publicly available cohorts (**Supplementary Table 1**). T1D case cohorts were matched to control cohorts based on country of origin and genotype array where possible as previously described^3^. We applied the HRC imputation preparation program (version 4.2.9, https://www.well.ox.ac.uk/~wrayner/tools/) and used PLINK^50^ (version 1.90) to perform quality control prior to imputation to remove variants with MAF <1%, missing genotypes >5%, in violation of Hardy-Weinberg equilibrium (HWE *P* <1×10^−5^ in control cohorts and HWE *P*<1×10^−10^ in case cohorts), difference in allele frequency > 0.2 compared to HRC r1.1 reference panel^51^, and variants with allele ambiguity.^3,50^. We then imputed genotypes for all samples into the TOPMed v2 and Michigan Multi-ethnic HLA reference panels using the respective imputation servers^52,53^. We additionally used SNP2HLA to impute genotypes into the specialized European HLA TDGC reference panel^54^. In the genome-wide imputation, we removed variants with an imputation accuracy score of r^2^<0.3. In the HLA imputed imputation, we removed variants with an imputation accuracy of r^2^<0.5 and a standard deviation in control allele frequency >0.055 across cohorts. Variants that passed QC filters in all cohorts were tested for association.

### Association testing and meta-analysis

We defined distinct DR3/DR4 and non-DR3/DR4 groups based on HLA-DR and HLA-DQ haplotype status using four-digit HLA alleles imputed from the T1DGC reference panel with SNP2HLA. DR3 was identified by the presence of HLA-DRB1*03:01-DQB1*02:01, while DR4 was identified by the presence of HLA-DRB1*04:01/02/04/05/08-DQB1*03:02/04/02:02^47^. We excluded 23 individuals who were identified as non-DR3/DR4 via SNP2HLA alleles, but had DR3 or DR4 tag SNPs^55,56^. From the non-DR3/DR4 risk group, 100 cases and 300 control samples we removed prior to performing association analyses. In both non-DR3/DR4 and DR3/DR4 groups, we tested variants for T1D case and control association using firth bias corrected logistic regression in EPACTS^57^. We tested variants with MAF>1% for association using sex and the first 4 genotype PCs as covariates. Summary statistics were combined across cohorts in a fixed effects inverse variance weighted meta-analysis using METAL^58^. The genomic inflation (lambda GC) was 1.069 for non-DR3/DR4 and 1.098 for DR3/DR4. We used LDSC^32^ to test for heritability in the summary statistics outside of the MHC for each group using a population prevalence of 1% for each group, and a sample prevalence of 51% for DR3/DR4 and 10% for non-DR3/DR4.

### Fine-mapping of independent signals

We identified 1 Mb regions around the lead variants of all genome-wide significant loci. We performed conditional analysis at each locus using stepwise analysis iteratively including the most significant variant in the regression model and re-performing the meta-analysis until no significant variants remained. In conditional analysis we used a locus wide significance threshold of p< 1×10^-5^. We then performed Bayesian fine mapping to create credible sets of likely causal variants for each signal^59^. From the summary statistics, effect size and standard error were used to calculate the approximate Bayes factor (BF) for each variant in r^2^>0.1 with the lead variant. We calculated the probability of association (PPA) for each variant by dividing the BF by the total sum of BFs. We then created 99% credible sets by including variants in descending order of PPA until the cumulative posterior probability was at least 99% (**Supplementary Table 12**).

### ATAC-seq peak calling

ATAC-seq data for 175 samples from 20 different immune cell types (Bulk B, Mem B, Naïve B, Effector CD4pos T, Follicular T Helper, Memory Teffs, Memory Tregs, Naïve Teffs, Th1 precursors, Regulatory T, Th2 precursors, Th17 precursors, Naïve Tregs, Effector memory CD8pos T, Naïve CD8 T, Central memory CD8pos T, CD8pos T, Gamma delta T, Monocytes, Mature NK) at resting and stimulated conditions were obtained from the NCBI GEO database at the accession GSE118189^34^ and processed to generate peak coordinates. Reads were aligned using STAR^60^ to hg19 and duplicate reads, reads mapping to blacklisted regions from ENCODE, and read with mapping quality Q<30 were filtered. Peak calling was performed using MACS2^61^ on BAM files further filtered for read pairs with insert size no larger than 140 bp (macs2 callpeak --nomodel --nolambda --keep-dup all --call-summits -f BAMPE -g hs –q 0.01), combining individual samples for each cell type and treatment, for a total of 40 distinct peak sets. We then used bedtools multiinter to obtain one consensus set of peaks and featureCounts^62^ to obtain the peak x read counts in each cell-type.

### GWAS enrichment analysis

For each group, we performed partitioned heritability LD-score regression to estimate genome wide enrichment in stimulated and unstimulated immune cell accessible chromatin generated above^32–34^. We used the summary statistics for each group excluding the MHC locus and formatted it for input to LD score regression using the munge_sumstats.py script. We generated binary annotations from each accessible chromatin bed file and computed cell specific LD enrichment scores for each risk cohort using the version 2.2 1000G baseline model. We then corrected for multiple tests in each group across all cell types using false discovery rate (FDR) and considered FDR<0.10 significant. Additionally, we performed gene set enrichment in GO, KEGG and REACTOME pathways with the summary statistics for each group using MAGMA^63^ with default parameters and reported the strongest enrichments using uncorrected p-values.

### Testing differences in T1D risk between groups

We tested for differences in T1D effect between non-DR3/DR4 and DR3/DR4 using merged PLINK files of Michigan HLA and TOPMed imputed variants for all 29,723 samples. We tested for heterogeneity in marginal effects on T1D using Breslow-Day (BD) tests with the “-bd” flag in PLINK^50^. We performed BD tests for lead variants for six non-HLA loci identified in non-DR3/DR4 samples, and lead variants for 88 previously known T1D risk loci^3^. At MHC and other loci with multiple signals, we tested for heterogeneity in effects on T1D conditional or other known variants. We generated regression models with PLINK using the “-glm interaction firth” flag including sex, the first 4 genotype PCs, DR3/DR4 status, and additional variants as covariates and evaluated the interaction with DR3/DR4 status. For the 12 MHC signals identified in non-DR3/DR4 samples, we conditioned on preceding lead variants from the stepwise regression. For the *IFIH1* and *PTPN2* loci, we conditioned on lead variants for all other known signals at the locus^50^. For the HLA locus, we conditioned on HLA-DRB1*03:01, HLA-DQB1*02:01, HLA-DRB1*04:01, HLA-DRB1*04:02, HLA-DRB1*04:04, HLA-DRB1*04:05, HLA-DRB1*04:08, HLA-DQB1*03:02, and HLA-DQB1*03:04 to examine heterogeneity in 40 known two-digit HLA risk alleles independent of these 9 DR3 and DR4 alleles. To then obtain the conditional effect of each variant within non-DR3/DR4 or DR3/DR4, we performed logistic regression separately for each group using EPACTS including sex, 4 genotype PCs, and the same variants from the interaction tests above.

### Generation and statistical analysis of the GRSs

We calculated GRS1 by using SNP2HLA imputed genotypes to define DR3 and DR4 tag SNPs and TOPMed imputed genotypes for the remaining 28 signals^55^. We calculated GRS2 by using TOPMED variants where possible (60 variants), Michigan HLA for rs116522341, rs1281934, and the Michigan HLA proxies DQB1*06:02, B*18:01, DPB1*03:01, rs1611547, and rs114170382, for rs17843689, rs371250843, rs559242105, rs144530872, and rs149663102 respectively. We excluded individuals with more than 2 HLA-DR/DQ calls, for GRS2, in line with the published methods^35^. For each of the signals identified in the non-DR3/DR4 group, we generated an 18-varaint GRS by summation of all signals weighted by the beta (*B*) for each effect allele (*X*) (**Equation 1**, **Supplementary Table 12**).

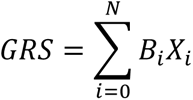

We additionally leveraged 82 non-HLA T1D risk loci from the largest T1D GWAS conducted to date^3^ weighted by non-DR3/DR4 marginal effects to generate a 100-variant combined GRS. We determined the ability of GRS2, the 18-variant GRS, and 100-variant GRS to distinguish non-DR3/DR4 T1D from either all control samples (DR3/DR4 and non-DR3/DR4) or non-DR3/DR4 control samples. We tested the ability of each GRS to discriminate T1D and non-diabetes using the area under the curve (AUC) of the receiver operator characteristic (ROC) statistics^64^. We calculated the difference between AUCs of each GRS using the deLong test and calculated the diagnostic criteria using the Youden Index. We then generated percentiles for how many non-DR3/DR4 T1D or control individuals fall at various GRS score thresholds. We calculated sensitivity at each GRS score as TP/ (TP + FN) and specificity as TN/(TN+FP)^65^. We further tested the ability to differentiate T1D from non-disease in the 100-variant non-DR3/DR4 T1D GRS using an independent test group of 100 cases and 300 control samples removed prior to association analyses. We also compared the ability of each GRS to differentiate T1D from T2D using 1,999 T2D individuals from the WTCCC study^66^. After imputing the T2D genotypes into TOPMed and Michigan HLA reference panels, we calculated the scores for each GRS as described above and derived ROC statistics comparing T1D to T2D. We additionally validated the performance of the 100-variant non-DR3/DR4 GRS to distinguish T1D from T2D using the 100 T1D case test group and all 1,999 T2D samples.

## Data Availability

All data produced in the present study are available upon reasonable request to the authors

## Supplement

**Supplementary Fig. 1.**
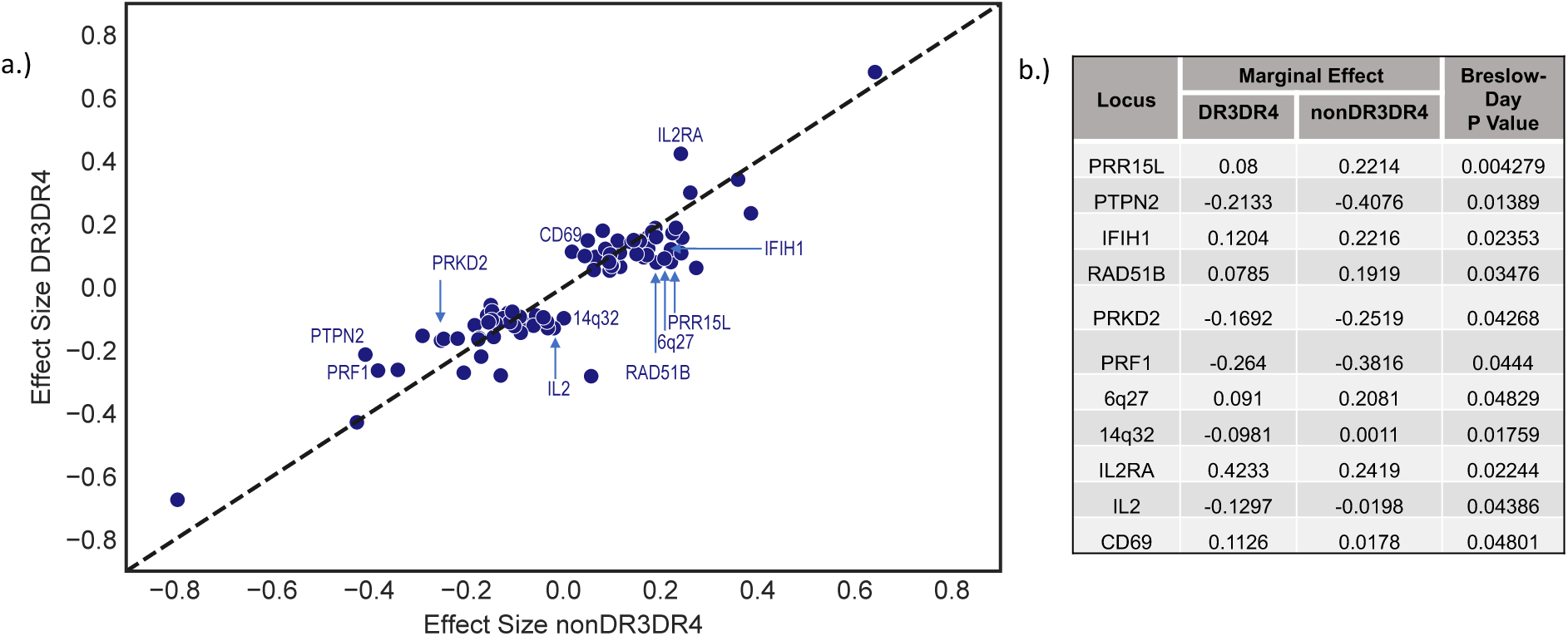
**a.** The T1D effect size of lead variants in non-DR3/DR4 and DR3/DR4 at primary signals for 88 known T1D loci. Selected loci with variants that have the largest differences in T1D effect are annotated. **b.** Table containing the locus and T1D effect size for non-DR3/DR4 and DR3/DR4 and the p-value for heterogeneity in effect using a Breslow Day test for 11 lead variants with p <0.05.

**Supplementary Fig. 2.**
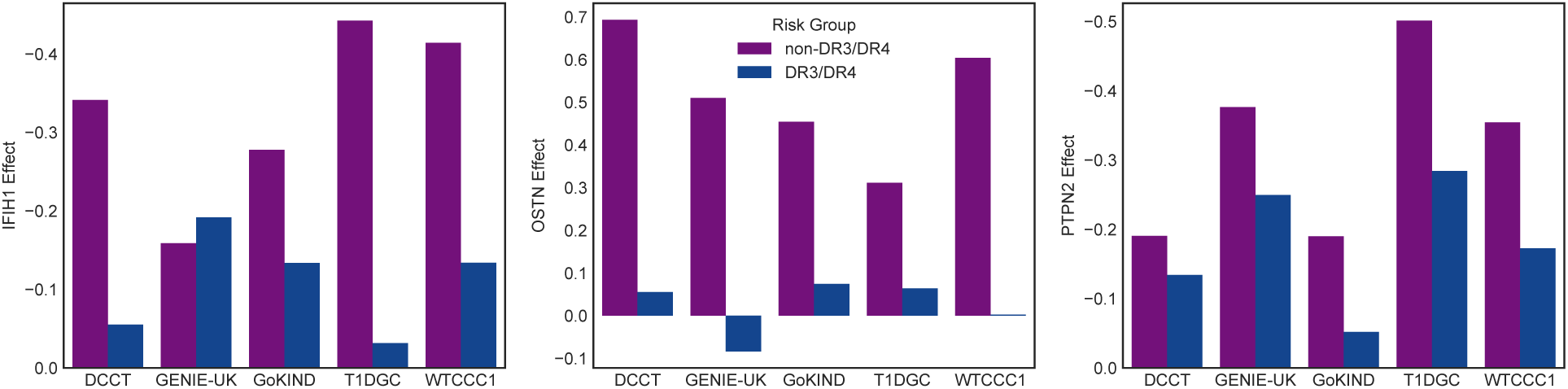
Effects for discovery non-MHC non-DR3/DR4 signals. The effects of lead variants at the *IFIH1, OSTN* and *PTPN2* loci on T1D risk in non-DR3/DR4 and DR3/DR4 stratified by individual cohort included in this study.

**Supplementary Fig. 3.**
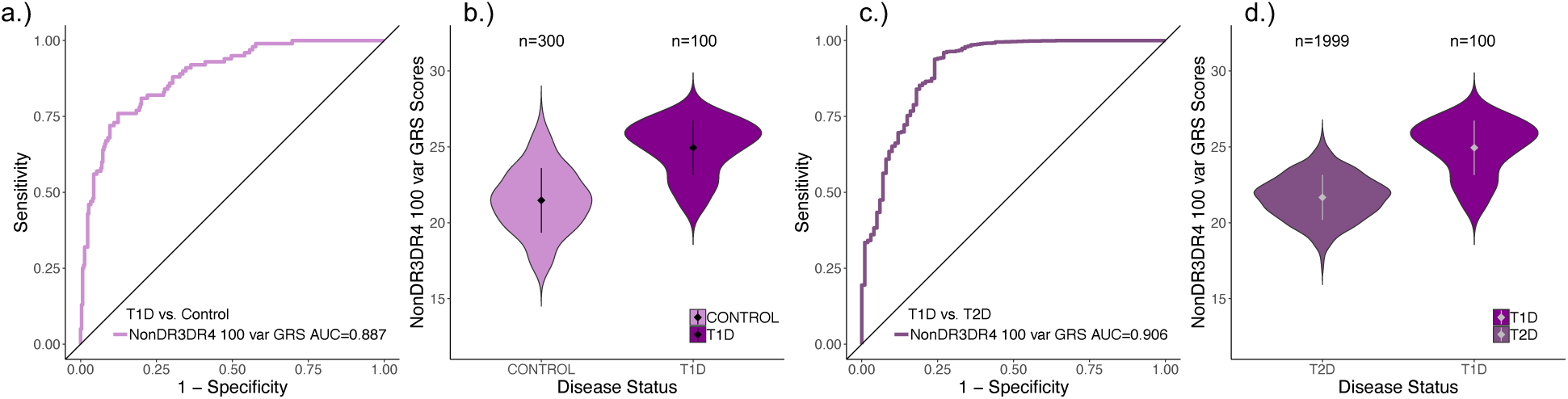
ROC curves showing the ability of GRSs to differentiate T1D from non-disease and T1D from T2D in an independent test population of non-DR3/DR4 T1D and corresponding violin plots. **a.** The AUC is shown for the 100-variant non-DR3/DR4 T1D GRS in non-DR3/DR4 T1D and non-disease and **b.** violin plot for scores in 100 T1D cases and 300 controls from the test group. **c.** The AUC is shown for the combined 100-variant non-DR3/DR4 T1D GRS in non-Dr3/DR4 T1D and all T2D. **d.** violin plot for scores in 100 T1D cases from the test group and 1999 T2D individuals.

